# Angiotensin Converting Enzyme Inhibitors and Angiotensin Receptor Blockers and Outcome of COVID-19 : A Systematic Review and Meta-analysis

**DOI:** 10.1101/2020.05.06.20093260

**Authors:** Aref A. Bin Abdulhak, Tarek Kashour, Anas Noman, Haytham Tlayjeh, Ala Mohsen, Mouaz H. Al-Mallah, Imad M. Tleyjeh

## Abstract

**Importance:** There is conflicting evidence about the role of angiotensin converting enzyme inhibitors (ACEIs)/angiotensin receptor blockers (ARBs) in the pathogenesis and outcome of patients infected with acute severe respiratory syndrome coronavirus 2 (SARS-CoV-2) and growing public concerns about their use during this pandemic.

**Objective:** We sought to systematically review the literature and perform a meta-analysis about prior use of ACEI/ARBs and outcome of COVID-19 patients.

**Data source:** We searched multiple data sources including PubMed, ClinicalTrial.org, and medrxiv.org from November 2019 through May 3, 2020.

**Study selection:** Any study that reported on the role of ACEIs / ARBs and outcome of COIVD-19 is eligible. Two authors independently reviewed eligible studies and extracted data into a prespecified data collection form.

**Data synthesis:** An inverse variance meta-analytic approach was used to pool adjusted odds ratios using a random effect model meta-analysis. I 2 test was used to assess in between studies heterogeneity. The Newcastle-Ottawa quality assessment scale was used to assess the quality of included studies.

**Main outcome and Measures:** The association between the prior use of ACEIs / ARBs and the mortality among SARS-CoV-2 infected patients was assessed using pooled OR and 95% confidence interval. For studies that did not report adjusted effect estimates for mortality, we used their adjusted effect estimate of critical outcome to estimate another pooled OR for critical or fatal outcome and its 95% confidence interval.

**Results:** Nine studies were included in this systematic review. The studies included a total of 58,615 patients infected with SARS-CoV-2. Prior use of ACEIs and/or ARBs was associated with a significant reduction of inpatient mortality among SARS-CoV-2 infected patients, with a pooled adjusted OR from 4 studies of 0.33, 95% CI [0.22, 0.49] with no between studies heterogeneity (12=0%) and with a significant reduction of critical or fatal outcome, with a pooled adjusted OR from 5 studies of 0.32, 95% CI [0.22, 0.46] (12 =32%).

**Conclusion:** Our meta-analysis suggests that use of ACEIs/ARBs is associated with a decreased risk of death or critical outcome among SARS-CoV-2 infected patients. This finding is limited by the observational nature of the included studies. However, it provides a reassurance to the public not to stop prescribed ACEIs/ARBs due to fear of severe COVID-19. It also calls upon investigators and ethics committees to reconsider the ongoing randomized trials of discontinuation of these drugs.

## Introduction

The Coronavirus disease 2019 (COVID-19) caused by the Severe Acute Respiratory Syndrome Coronavirus 2 (SARS-CoV-2) emerged in December 2019 in Wuhan China and since then spread to the rest of the world. By May 02, 2020, the disease has been reported in 215 countries with over 3 million confirmed cases and over 230,000 deaths^1^.

Several studies have demonstrated that subjects with underlying cardiovascular diseases (CVDs) represent a significant proportion of patients with symptomatic COVID-19 disease and were among the highest risk individuals for severe disease and worse prognosis with 5-10 fold increase in mortality^2-5^. Additionally, several studies revealed that COVID-19 disease is associated with a number of cardiovascular complications including, heart failure, cardiac arrhythmias, pericarditis, myocarditis and vasculitis and Up to 28% of COVID-19 patients experience elevated troponin levels^6-13^.

The newly emerged SARS-CoV-is a betacoronavirus, which is related to the SARS-CoV virus that caused SARS outbreak in 2003 with 80% genomic homology^14^. A few studies established that SARS-CoV-2 invades human cells through binding of its spike S-protein to angiotensin converting enzyme 2 (ACE2) with the help of the host TMPRESS2 membrane serine protease that primes the virus Spike (S) protein facilitating its cell entry^15-17^. The 3D structure of SARS-Cov-2 binding site indicate that it has improved binding stability and it is estimated that its binding affinity to ACE2 is 10-15 times higher than that of SARS-CoV^18-19^.

Angiotensin converting enzyme 2 (ACE2) is a type-1 transmembarne glycoprotein that is expressed mainly in the testis, intestine, adipose tissue, cardiovascular system and the lung. It catalyzes the conversion of angiotensin-II to angiotensin (1-7) peptide, which has several cardiovascular protective functions such as vasodilatation, anti-proliferative, anti-fibrotic and anti-inflammatory effects ^20^. ACE2 also exists as a soluble form and serum levels of ACE2 were found to be elevated in patients with hypertension and heart failure^21-22^. Moreover, ACE2 has immune modulating effects as shown in several animal models of acute lung injury including SARS model. Furthermore, in a phase-II human trial, recombinant ACE2 infusion significantly reduced angiotensin-II and IL-6 levels in patients with acute respiratory distress syndrome (ARDS), and observed that non-survivors had higher angiotensin-II in comparison to survivors^24^. Interestingly, serum angiotensin-II was reported to be significantly increased in patients with COVID-19 disease and correlated positively with lung injury and viral titers^25^.

Another confounding issue is the fact that several animal studies illustrated that ACE inhibitors (ACEIs) and angiotensin receptor blockers (ARBs) upregulate ACE2 expression, which raises concerns that these agents, which are widely used to treat hypertension and other cardiovascular conditions may increase susceptibility to SARS-CoV-2 infection^26-28^.

These observations led to the speculation that the patients using ACEIs or ARBs might be more susceptible to SARS-CoV-2 infection and severe COVID-19 disease^29^. This created a major controversy within the scientific community because of the lack of supporting human data; and hence prompted several scientific societies to issue statements regarding the use of ACEIs and ARBs in patients with COVID-19 disease warning against discontinuation of such medications in the absence of supporting convincing evidence.

Moreover, at least 5 randomized trials in multiple countries plans to enroll COVID-19 patients on prior ACEI/ARBs to investigate whether stopping these drugs would affect COVID-19 outcome.

Therefore, herein we aimed to systematically review the exiting literature and perform a meta-analysis of the available studies examining the role of ACEIs/ARBs in the outcome of COVID-19.

## Methods

A comprehensive systematic review and meta-analysis was conducted according to Preferred Reporting Items for Systematic Reviews and Meta-analyses (PRISMA)^30^ and Meta-Analysis of Observational Studies in Epidemiology guidelines for reporting systematic review and meta-analysis of observational studies guidelines^31^.

### Study selection

Any study that reported the association between the prior use of angiotensin converting enzyme inhibitor and or angiotensin receptor blocker and the outcomes of infected patients with COVID-19 is eligible. Two authors independently (AB and IT) examined eligible studies.

### Search Strategy

Medline was searched from November 2019 through May 3, 2020, without language restriction for eligible studies. Pubmed search keys included (“ coronavirus “ OR “covid-19” OR “SARS” AND “ Angiotensin” OR “Angiotensins”). ClinicalTrial.org and medrxiv.org were also searched using similar PubMed search terms.

### Data collection and quality assessment

A data collection form was filled by the two authors independently (AB and AM) and reviewed by the senior author (IT). The form contains information about study’s population, study’s characteristics, exposure to ACE and or ARB, and outcomes including death and severity of infection with COVID-19.

The eligible studies were assessed according to the Newcastle–Ottawa quality assessment scale (NOS)^32^. NOS scale rates observational studies based on 3 parameters: selection, comparability between the exposed and unexposed groups, and exposure/outcome assessment. It assigns a maximum of 4 stars for selection, 2 stars for comparability, and 3 stars for exposure/outcome assessment.

Studies with less than 5 stars were considered low quality, 5–7 stars moderate quality, and more than 7 stars high quality. Two reviewers (AN and HT) assessed the quality of the included studies.

### Analytical method

An inversed variance meta-analytic approach was used to pool study’s adjusted odds (OR) ratio or hazards ratio (HR) with its 95% confidence interval (CI) into a random effects model meta-analysis. The two outcomes that were considered for this analysis were death as the primary outcome of this meta-analysis and when adjusted mortality estimates were not reported, we used critical illness as a surrogate of a combined (critical/fatal) outcome resulting from the infection with SARS-CoV-2. We used adjusted ORs or HRs from propensity score matching whenever available or multivariate analysis to pool the overall effect estimate. We collected data for ACEI and ARB effect estimates separately whenever reported in individual studies. We only included adjusted effect estimates because of the observational design of all included studies.

Heterogeneity among the included studies was assessed using χ^2^ and I^2^ tests. The I^2^ statistic describes the proportion of variation in treatment estimate that is not related to sampling error^33^. A value of zero indicates no heterogeneity, 25%–49% low, 50%–74% moderate, and 75% a high degree of heterogeneity. No formal assessment of publication bias was performed because of the low number of studies as per Cochrane guidelines.

Two-sided P value of < 0.05 was considered statistically significant for all included analyses. The statistical software Review Manager, version 5.3.5 (The Nordic Cochrane Center, The Cochrane Collaboration, 2018, Copenhagen, Denmark) was used for all analyses.

## Results

### Search strategy and general characteristics of included studies

Figure 1 showed the result of our search strategy. A total of 9 studies^34-42^ (8 cohort studies and one population-based case control study) met our eligibility criteria. Table 1 shows the general characteristics of included studies. There were four studies from China, one study from the USA, one from United Kingdom, one from Italy, and one study was an international collaboration. A population of 58,615 patients with COVID-19 were included in this systematic review and meta-analysis. Seven out of 9 included studies reported data on our outcome of interest (mortality) but only 4^34, 35, 37, 41^ of them reported adjusted OR. Only one^36^ paper reported adjusted OR for severity of the illness (for ARBs use only) but did not report on the mortality; thus, it was included in the combined critical/fatal outcome. Therefore, the outcome of interest (combined critical / fatal outcome) included only 5 studies. Four out of 9 studies reported combined ACEIs with ARBs as the primary exposure and the rest of the studies reported data separately on ACE and ARB. Table 2 summarizes the analytic approach and result of qualityassessment of the included studies. There were only few reported data to perform a separate analysis involving ARBs only.

**Figure 1:**
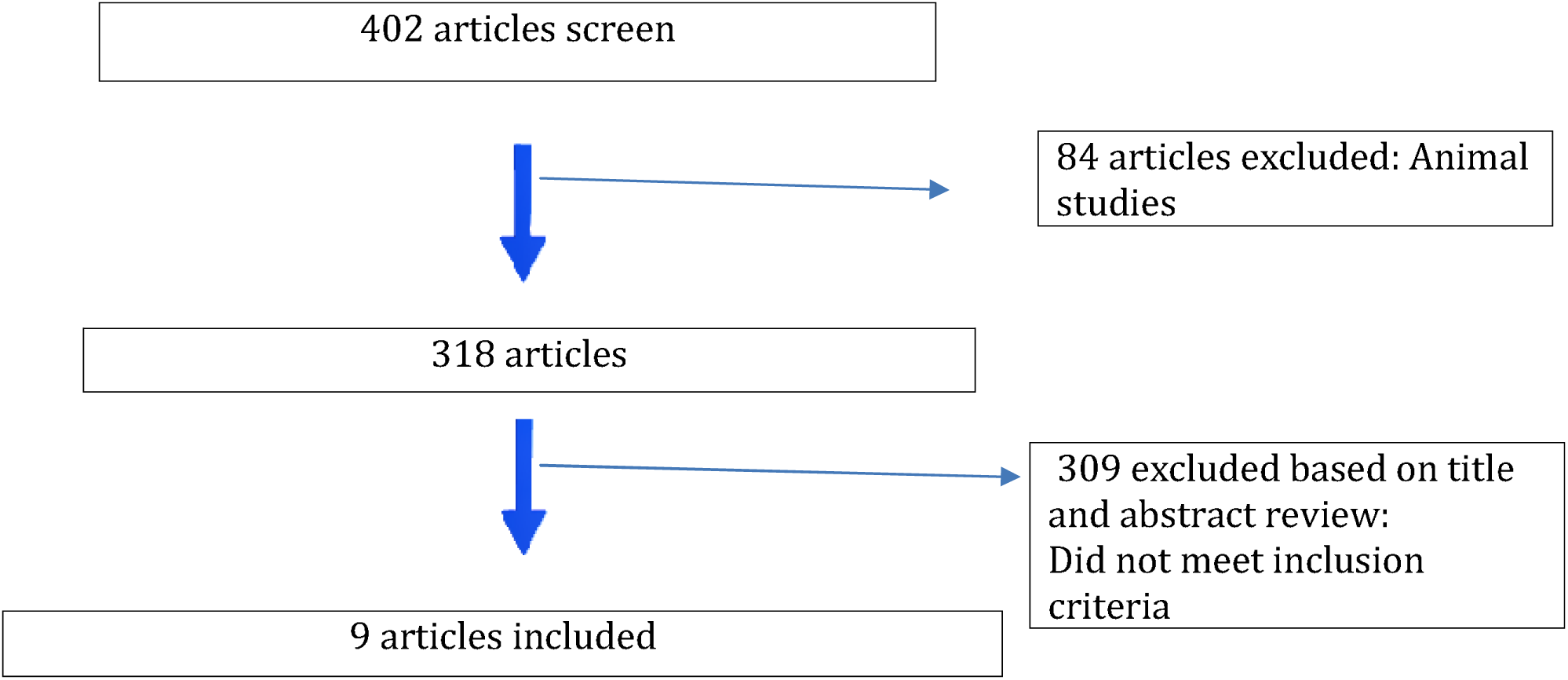
Flow diagram of included studies

**Table 1:**
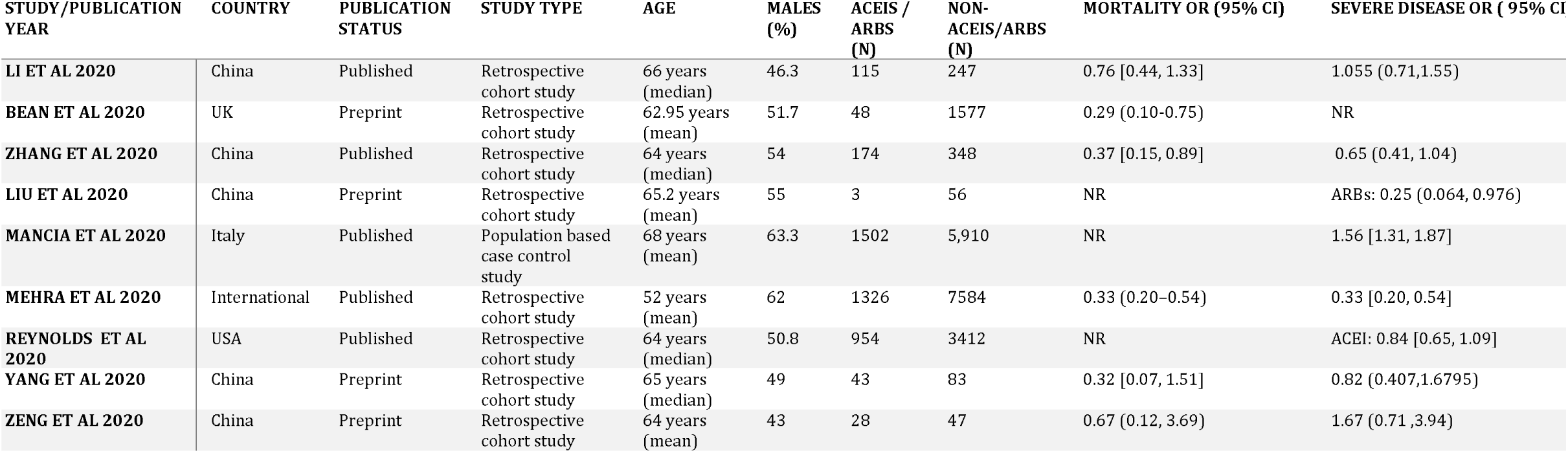
General characteristics of included studies. ACEIs: angiotensin converting enzyme inhibitors, ARBs: angiotensin receptor blockers, ADRS: adult respiratory distress syndrome, CL: confidence interval,NR: not reported, OR: odds ratio

**Table 2:**
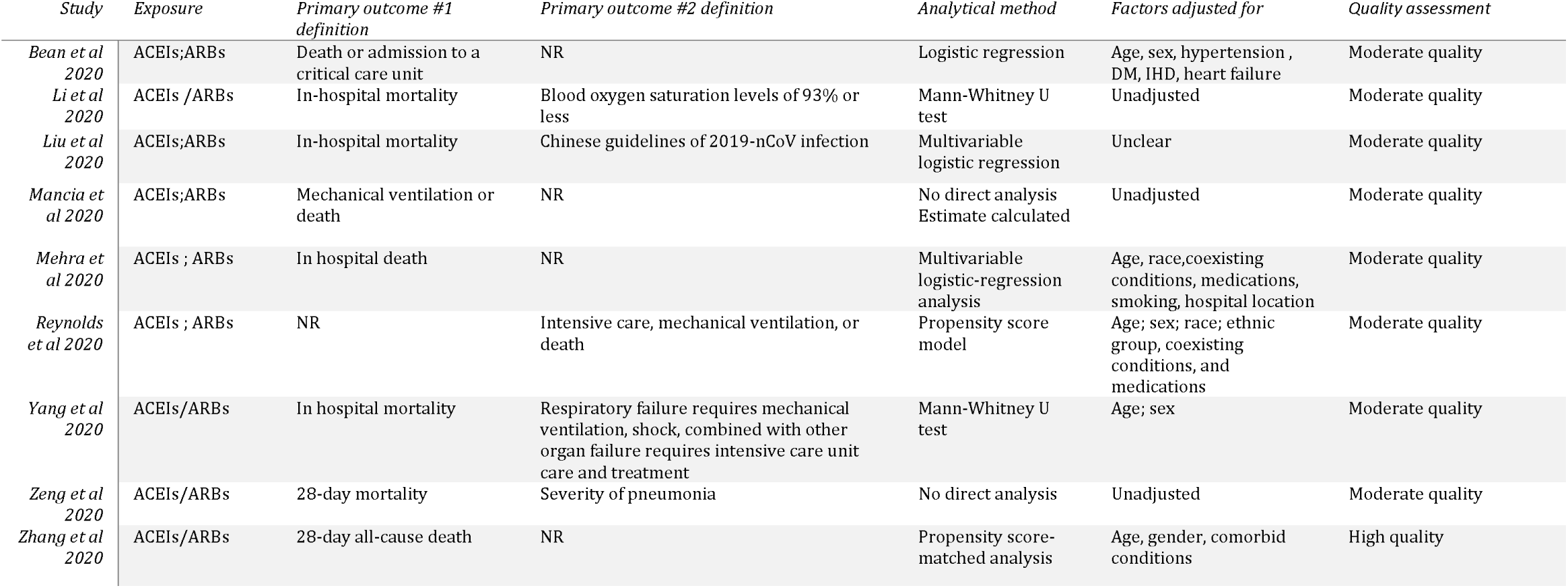
Outcomes and analytic approaches of included studies.

**Figure 2:**
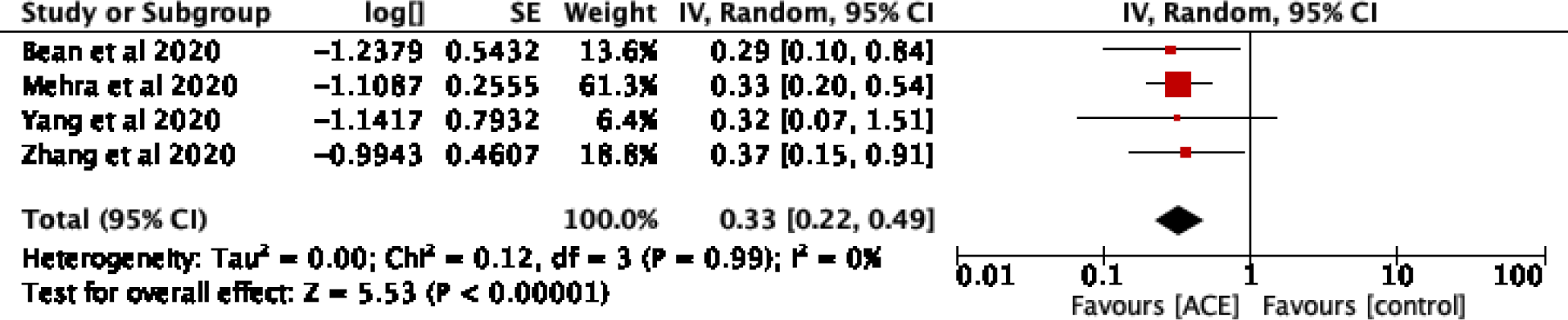
Random effect model meta-analysis of the adjusted effect estimates of the association between ACEIs/ARBs user and death from COVID-19

**Figure 3:**
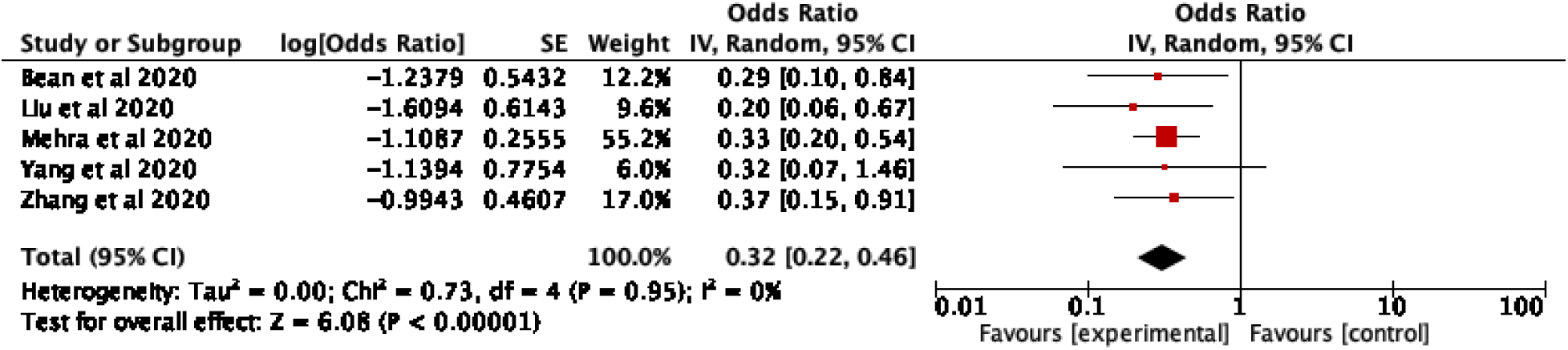
Random effect model meta-analysis of the adjusted effect estimates of the association between ACEIs/ARBs user and critical or fatal outcome from COVID-19

All studies included in the 2 analyses were of moderate quality but one study^34^ scored high as adjudicated by NOS scale as shown in Table 2.

### Meta-analysis

Prior use of ACEIs and/or ARBs was associated with a significant reduction of inpatient mortality among SARS-CoV-2 infected patients, with a pooled adjusted OR from 4 studies of 0.33, 95% CI [0.22, 0.49] with no between studies heterogeneity (I^2^=0%) and with a significant reduction of critical or fatal outcome, with a pooled adjusted OR from 5 studies of 0.32, 95% CI [0.22, 0.46] (I^2^=32%).

## Discussion

Findings from our systematic review and meta-analysis suggest that prior ACEIs or ARBs use may be associated with a lower risk of fatal or critical outcome in hospitalized patients with COVID-19 infection.

Although the exact mechanism(s) behind the observed association remains unknown, it could be attributed to several factors including cardioprotective effects of ACEIs and ARBs or their direct effect on lung inflammation, viral replication, cytokine production and/or their effects on the interaction of the virus with the renin-angiotensin-aldosterone system.

Cardiovascular complications are common in patients with pneumonia in general and in COVID-19 patients^43^. A recent systematic review and meta-analysis of 39 observational studies between 1984 to 2019, involving 92,188 patients, reported cardiac complications in 13.9% (95% confidence interval (CI) 9.6–18.9), acute coronary syndromes in 4.5% (95% CI 2.9–6.5), heart failure in 9.2% (95% CI 6.7–12.2), arrhythmias in 7.2% (95% CI 5.6–9.0) and stroke in 0.71% (95% CI 0.1–3.9) of pooled inpatients data^44^.

Moreover, cardiovascular disease risk factors are common in patients admitted with COVID-19 and this could potentially explain the observed beneficial association between prior use of ACEIs/ARBS and outcomes. In a recent meta-analysis of 7 studies including 1576 COVID-19 patients, the most prevalent comorbidities were hypertension (21.1%, 95% CI: 13.0-27.2%) and diabetes (9.7%, 95% CI: 7.2-12.2%), followed by cardiovascular disease (8.4%, 95% CI: 3.8-13.8%) and respiratory system disease (1.5%, 95% CI: 0.9-2.1%)^45^.

Our findings are concordant with previous studies of ACEIs and ARBs in community acquired pneumonia in general. In a previous systematic review and meta-analysis investigators reported that, compared with control treatments, both ACEIs (seven studies: odds ratio 0.73, 0.58 to 0.92; I^2^=51%) and ARBs (one randomized controlled trial: 0.63, 0.40 to 1.00) were associated with a decrease in the pneumonia related mortality^46^.

In another single center cohort of patients hospitalized with acute respiratory illness who tested positive for respiratory viruses by PCR, and developed concomitant pneumonia, the continued use of ACEI was associated with a decreased risk of mortality or intubation. Furthermore, those who were on ACEI prior to admission and subsequently discontinued the medication had a higher mortality than those not on an ACEI prior to admission. The authors speculated that the ACEI were discontinued in patients with shock or acute kidney injury, thus representing more severe viral pneumonia than those in whom it was continued throughout their hospital stay^47^.

In another large propensity matched cohort of around 23000 patients aged ≥65 years hospitalized with pneumonia from the Department of Veterans Affairs, prior ACEI use (OR, 0.88; 95% CI, .80–97) and inpatient ACEI use (OR, 0.58; 95% CI, .48–.69) was associated with decreased mortality. Prior ARB use (OR, 0.73; 95% CI, .58–.92) and inpatient ARB use (OR, 0.47; 95% CI, .30–.72) was also associated with decreased mortality^48^.

Mechanistically, numerous studies investigated the role of RAAS in the pathogenesis of acute lung injury in different animal models. Indeed, several of them reported increased ACE activity and elevated angiotensin-II levels in acute lung injury induced by sepsis, lipopolysaccharide, acute pancreatitis and ventilator induced injury. In addition, these studies demonstrated that treatment with either ACEIs or ARBs ameliorated acute lung injury in these models ^49-52^ ACEIs and ARBs seem to exert their protective effect through modulation of inflammatory and immune responses. They led to reduction in inflammatory cytokines, TNF-alpha, IL-1beta and IL-6^49^, inhibition of dendritic cell maturation and Th1 and Th17 cell polarization responses^50^, and suppression of Rho/ROCK pathway^51^. These inflammatory and immune modulating effects of ACEIs and ARBs may have clinical relevance in the context of COVID-19 disease. In a recent study, Meng et al compared hypertensive patients on ACEIs/ARBs with those on other medications and found that patients on ACEIs/ARBs had higher CD3+ and CD+ T cells and a trend of lower IL-6. They also observed that although there was no difference in the viral load between the two groups on admission, patients on ACEIs/ARBs had lower peak viral load^53^.

The other axis of the RAAS is comprised of ACE2 with its main effect is to counterbalances the deleterious effects of angiotensin-II that is produced by ACE. ACE2 promotes the conversion of angiotensin-I to angiotensin-(1-9) peptide and angiotensin-II to angiotensin-(1-7) peptide, which interacts with the Mas receptor and counteracts the effects of angiotensin-II^20^. Interestingly, angiotensin-II can reduce cell surface expression of ACE2 through enhanced cleavage of ACE2 by TNF-alpha converting enzyme (TACE/ADAM-17), establishing a feedback mechanism where by angiotensin-II promotes the loss of its negative regulator^54^ The interaction of angiotensin-II and ACE2 in the context of viral induced acute lung injury have been also well established in the mouse model of SARS where it has been shown that SARS-CoV infection or injection of SARS-CoV Spike protein reduced expression of ACE2 and that treatment with recombinant ACE2 or blocking angiotensin-renin pathway protect mice from lung injury^55^. Similar protective effect by recombinant ACE2 were observed against lethal infection with avian influenza A H5N1 virus^56^. Several other studies in animal models of acute respiratory distress syndrome (ARDS) demonstrated that treatment with recombinant ACE2 or angiotensin-(1-7) peptide attenuate lung injury and improve oxygenation ^57-59^. Relevant to these animal observations, Reddy et al^60^ reported recently in a pilot study of patients with ARDS that plasma angiotensin-(1-7)/ angiotensin-(1-10)ratio and angiotensin-(1-9)/ angiotensin-(1-10) ratio correlated with survival indicating that ARDS survivors had higher ACE2 activity implying possible protective role of ACE2/angiotensin-(1-7) axis in ARDS. Human experience with recombinant ACE2 therapy in ARDS is very limited ^24^. However, ongoing clinical trials with recombinant ACE2 in COVID-19 disease will shed light on the utility of such therapy in acute lung injury in humans.

Our inferences from this meta-analysis are, however, weakened by limitations inherent to the meta-analysis and the individual studies. Given the observational design of included studies and retrospective data collection, the possibility that the observed association between ACEI/ARB use and COVID-19 outcome was affected by bias or confounding should be considered. For example, comorbidities and severity of illness are known confounders that are associated with ACEI/ARB intake and mortality and the healthy user effect could theoretically explain a potential role for ACEI/ARB in infection outcome. However, the opposite could also be the case where the sicker patients have more exposure to ACEI/ARB because patients with hypertension and cardiovascular risk factors have higher mortality from COVID. Moreover, the combined effect estimate from the studies that used regression models to adjust for possible confounders including age, comorbidities, and severity of illness—although the models were not uniform— supports our conclusions. As with any observational study, residual confounding cannot be fully excluded.

Given our findings, it becomes ethically concerning to continue to enroll patients in the ongoing randomized trials that are testing the hypothesis that stopping/replacing chronic treatment with ACEI or ARB improves outcomes in symptomatic SARS-CoV2-infected patients. We are aware of at least 5 trials listed on clinicaltrias.gov. (Table 3).

Although our study suggests that there may be an association between prior ACEIs/ARBs intake and outcome of COVID-19 patients, it remains unknown whether de novo initiation of ACEIs/ARBs as an adjunctive therapy for COVID-19 will prove beneficial (Supplement table 1).

## Conclusions

Our meta-analysis suggests that prior use of ACEIs/ARBs is associated with a decreased risk of death or critical outcome among SARS-CoV-2 infected patients. This finding is limited by the observational nature of the included studies. However, it provides a reassurance to the public not to stop prescribed ACEIs/ARBs due to fear of severe COVID-19. It also calls upon investigators and ethics committees to reconsider the ongoing randomized trials of discontinuation of these drugs. Moreover, our findings may help encourage patients’ enrollment in the ongoing clinical trials evaluating the therapeutic role of de novo initiation of ACEIs/ARBs in SARS-CoV-2 infection.

## Data Availability

NA

